# Outcomes of Kidney Transplantation in Cystic Fibrosis

**DOI:** 10.1101/2024.09.19.24313959

**Authors:** Mirjana Stevanovic, Todd A MacKenzie, Asha Zimmerman, Martha L Graber

## Abstract

**Background:** Cystic fibrosis (CF) is a chronic multisystem disease with features including recurrent pulmonary infection, bronchiectasis, malnutrition, and alterations in drug metabolism which may discourage referral and listing for kidney transplantation. Although no specific renal phenotype is identified in CF, chronic kidney disease (CKD) occurs in young people and adults with CF with greater frequency than in the general population. The characteristics and outcomes of kidney transplantation in the CF population have not been previously described.

**Methods:** We used de-identified data supplied by the US Renal Data System (USRDS) to compare persons with end stage kidney disease (ESKD) with first kidney transplant, who did and did not have a diagnosis of CF. We compared demographic and clinical characteristics, mortality, patient survival, and death-censored graft survival using linear and logistic regression, multivariable logistic regression models, and Kaplan-Meier and log-rank tests with stratification/binning by propensity scores.

**Results:** Of those with dual diagnoses of CF and ESKD, half received a first kidney transplant. CF was independently associated with higher odds of receiving a living donor transplant. Those with a kidney transplant were younger than both those who did not have CF and those with CF without transplant and had higher odds of being female. Those with dual diagnoses of CF and ESKD had disproportionately lower odds of being identified as Black or African American relative to their representation in the population of all with CF and ESKD. Diabetes was more frequently a diagnosis and primary cause of ESKD in people with CF, but was approximately half as frequently the primary cause of ESKD as in those without CF. Complications of transplantation, particularly lung, were the second most frequent etiology of ESKD in people with CF. Overall survival with ESKD was 21.1 years for those with CF and 31.8 for those without CF. The survival benefit associated with transplant was 17.2 years for those with CF and 24.8 years for those without CF. Death censored median graft survival was significantly longer for those with CF, 20.4 vs 13.7 years.

**Conclusions:** CF is a chronic multisystem disease with features which may discourage referral and listing for kidney transplant. We used de-identified data from the USRDS datasets to describe the characteristics and outcomes of all unique individuals with the diagnosis of CF and first kidney transplant prevalent during the five-year period 2014 and 2018. Those with CF had similar odds to those without CF or receiving a first kidney transplant, and higher odds of receiving a living donor graft. Diabetes mellitus was significantly less frequently the primary cause of ESKD in those with CF and kidney transplant. Complications of solid organ transplant, particularly lung, were the second most frequent cause of ESKD in people with CF and kidney transplant. Median patient survival following the first kidney transplant was significantly shorter than for those who did not have CF, but was substantial, as was the survival advantage when compared to dialysis. Median graft survival was significantly longer for those with CF. It is our hope that this data will improve awareness of kidney disease in persons living with CF and their care teams, and encourage early referral to nephrology and transplant centers, and positive consideration of wait listing for kidney transplantation.

## Introduction

CF is a multi-system disease caused by mutations of the cystic fibrosis transduction regulator (CFTR), a transmembrane anion channel expressed in epithelial cells, including all tubular segments in the kidney^1^. CF is characterized by chronic pulmonary infection, bronchiectasis, pancreatic insufficiency, malnutrition, and alterations in drug metabolism^2,3^. CKD occurs with greater frequency in the population with CF than in the general population^4^, and may progress to end stage kidney disease (ESKD). Despite considerable improvements in health and longevity in recent decades^5^, the diagnosis of CF may be regarded as a contraindication to kidney transplantation. We used de-identified data provided by the US Renal Data System (USRDS)^6^ to report the characteristics and outcomes of kidney transplant patients with CF in the United States.

## Methods

CF is not a specified diagnosis on the ESKD Medical Evidence Report. We relied upon comparisons between all unique individuals who were listed in the USRDS Hospitalization dataset, carried a diagnosis of first kidney transplant status and did or did not have a diagnosis of CF. We compared characteristics using linear and logistic regression and performed univariable and age-adjusted comparisons for all characteristics. We compared the primary causes of ESKD between CF and non-CF using logistic regression. To test the association between CF and several outcomes of interest, we used multivariable logistic regression to control for patient characteristics including incidence age, sex, BMI, race or ethnicity, nephrologist care prior to ESKD, diabetes diagnosis, diabetes as the primary cause of ESKD, lung transplant status, complications of lung transplant as the primary cause of ESKD and living donor graft. We performed variable selection using forward and backward stepwise regression for each outcome. We chose the models with the lowest Akaike Information Criterion values. We compared patient and death-censored first kidney graft survival between CF and non-CF, and between transplant and non-transplant status in both CF and non-CF patients using Kaplan-Meier and log-rank tests after matching for age at entry into the registry, BMI, race, Hispanic ethnicity, diabetes and lung transplant status, diabetes and complications of lung transplant the primary causes of ESKD^7^.

Summary statistics for the racial and ethnic categories American Indian or Alaska Native, Asian, Native Hawaiian or Pacific Islander, and Other or Multiracial, were not included in these analyses because of small numbers of individuals within these categories with a diagnosis of cystic fibrosis. Some categories of primary cause of ESKD listed in the ESRD Medical Evidence Report were not included in these analyses.

## Results

All results refer to first kidney transplant. Unless otherwise indicated, univariable analyses are controlled for age at ESKD incidence.

### Characteristics and outcomes of those with first kidney transplant who did and did not have CF

#### Demographic characteristics and primary causes of ESKD

The USRDS Hospitalization dataset included 379 unique individuals with diagnoses of both CF and ESKD between January 1, 2014, and December 31, 2018, of whom 192 (50.6%) had first kidney transplant status. The Hospitalization dataset included 953,824 unique individuals without CF in the same period, of whom 249,012 (26.1%) had first kidney transplant status (p < 0.001).

Those with CF were younger at the time of ESKD incidence, 36.4 years vs 43.5 years (p < 0.001) and had lower BMI, 21.8 vs 27.7 (p < 0.001) (Table 1). They had higher odds of being female, 52.1% vs 40.2% (OR 1.57; 1.18, 2.09; p = 0.002), higher odds of being identified as White, 91.7% vs 67.2% (OR 5.58; 3.35, 9.32; p < 0.001), and lower odds of being identified as Black or African American, 6.3 % vs 25.7% (OR 0.18; 0.10, 0.33; p < 0.001), or of Hispanic ethnicity, 8.9% vs 14.4% (OR 0.52; 0.32, 0.86; p = 0.01). Those with CF had non-significantly greater odds of having been under the care of a nephrologist prior to ESKD incidence 57.8% vs 50.6% (OR 1.19; 0.75, 1.90; p = 0.47).

**Table 1:**
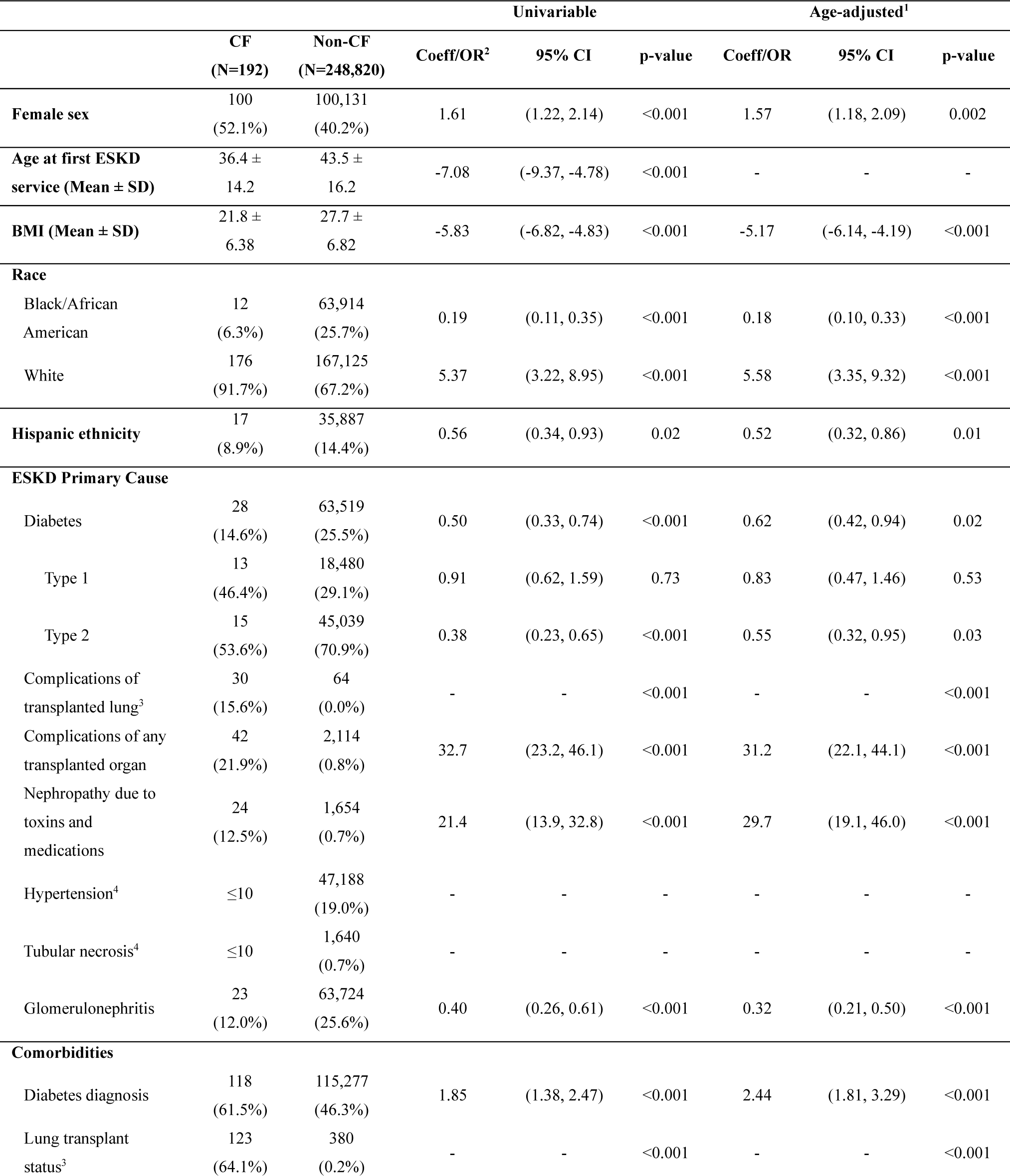
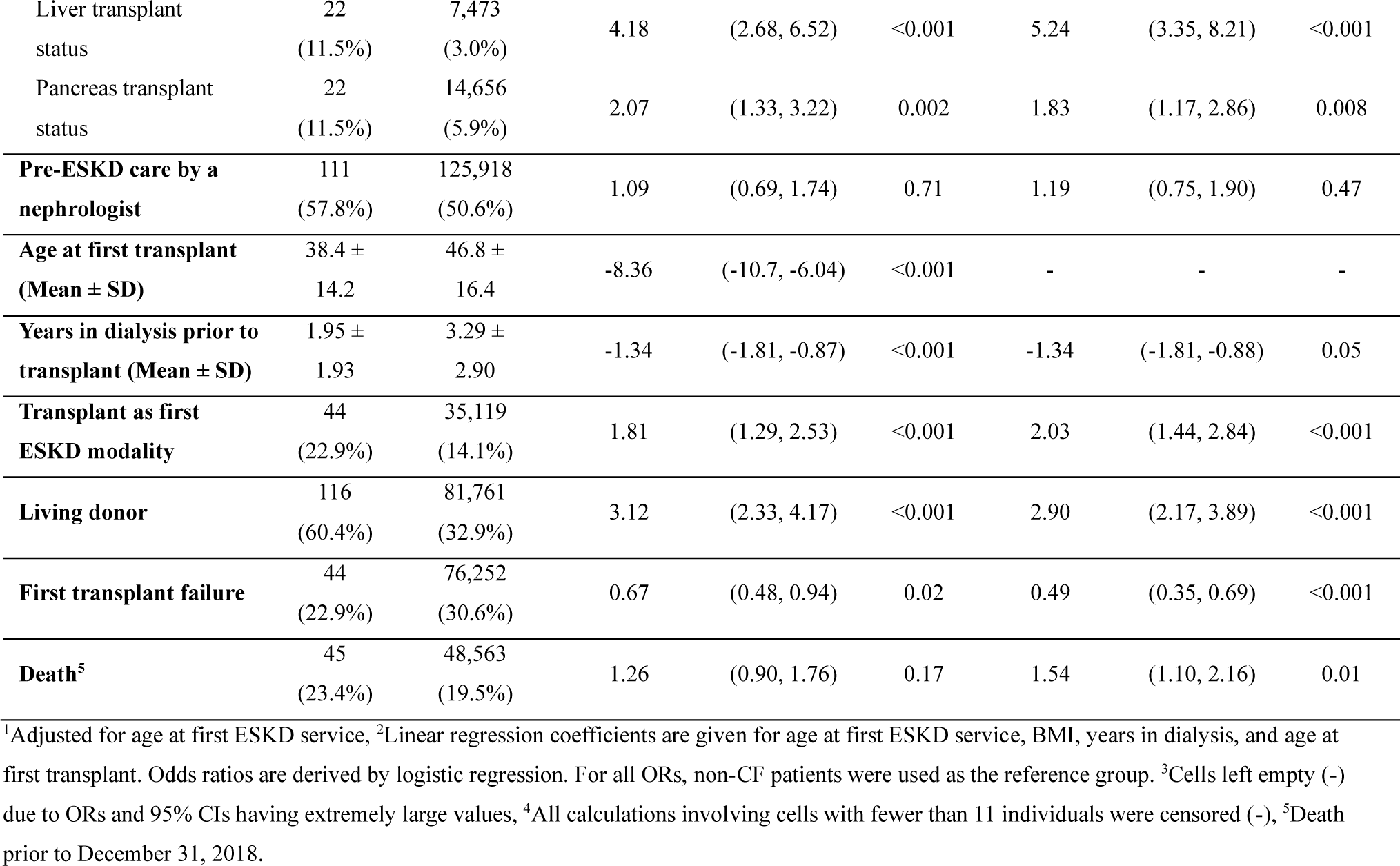
Characteristic of US kidney transplant patients with end stage kidney disease, with and without cystic fibrosis.

#### Primary causes of ESKD

In age-adjusted univariable analyses, persons with CF had higher odds of the primary cause of ESKD being complications of lung transplantation, 15.6% vs 0% (p < 0.001) and higher odds of complications of transplantation of lung or other organ being the primary cause of ESKD, 21.9% vs 0.8% (OR 31.2; 22.1, 44.1; p < 0.001). Of those with CF, 64.1% had a diagnosis of lung transplantation status, compared with 0.2% of those without CF. Liver transplantation status was documented in 11.5% of those with CF and in 3.0% of those without CF (OR 5.24; 3.35, 8.21; p < 0.001). Pancreas transplantation status was documented in 11.5% of those with CF and in 5.9% of those without CF (OR 1.83; 1.17, 2.86; p = 0.008). Nephropathy due to toxins and other medications was the primary cause of ESKD in 12.5% of those with CF and 0.7% of those without CF (OR 29.7; 19.1, 46.0; p < 0.001). Those with CF had lower odds than those without CF of diabetes as primary etiology of ESKD, 14.6% vs 25.5% (OR 0.62; 0.42, 0.94; p = 0.02). Diabetes was diagnosed in 61.5% of those with CF and in 46.3% of those without (OR 2.44; 1.81, 3.29; p < 0.001).

#### Transplant characteristics

People with CF were younger at the time of kidney transplant, 38.4 years, than those without CF, 46.8 years (p < 0.001), and had shorter time on dialysis prior to kidney transplant than those who did not have CF, 1.95 vs 3.29 years (p = 0.05). Those with CF had higher odds of having initiated ESKD care with a kidney transplant, 22.9% vs 14.1% (OR 2.03; 1.44, 2.84; p < 0.001). In multivariable analysis, CF was not independently associated with starting ESKD with a kidney transplant rather than dialysis (OR 1.18; 0.77, 1.82; p = 0.44). In age-adjusted univariable analysis, people with CF had higher odds of receiving a living donor graft, 60.4% vs 32.9% (OR 2.90; 2.17, 3.89; p < 0.001). In multivariable analysis, people with CF had higher odds of receiving a living donor graft (OR 1.75; 1.11, 2.76; p = 0.0163).

#### Patient and graft survival

In age-adjusted univariable analysis, those with CF had lower odds of graft failure, 22.9% vs 30.6% (OR 0.49; 0.35, 0.69; p < 0.001). In multivariable analysis controlling for incidence age, BMI, Black or African American, Asian, or White race, Hispanic ethnicity, pre-ESKD care by a nephrologist, years on dialysis prior to kidney transplant, dialysis type, diabetes diagnosis, diabetes as the primary etiology of ESKD, lung transplant status, and lung transplant as the primary cause of ESKD, CF was independently associated with lower odds of graft failure (OR 0.44; 0.27, 0.70; p < 0.001). In age-adjusted univariable analysis, those with CF had greater odds of death following kidney transplant prior to the end of the study period, 23.4% vs 19.5% (OR 1.54; 1.10, 2.16; p = 0.01). In multivariable analysis, CF was not independently associated with higher odds of death following kidney transplant (OR 1.29; 0.88, 1.89; p = 0.19). In Kaplan-Meier analysis with propensity score matching, median survival after kidney transplant was 18.4 years for those with CF and 27.4 years for those without CF (p = 0.0012) (Figure 1a). In Kaplan-Meier analysis with propensity score matching to compare those with CF with those without CF, median death-censored graft survival was 20.5 years for those with CF and 13.3 years for those without (p < 0.0001) (Figure 1b). In Kaplan-Meier analysis with propensity score matching, median survival for those without CF and with a kidney transplant was 30.5 years; median survival for those without CF who did not have a kidney transplant was 5.69 years (p < 0.0001).

**Figure 1:**
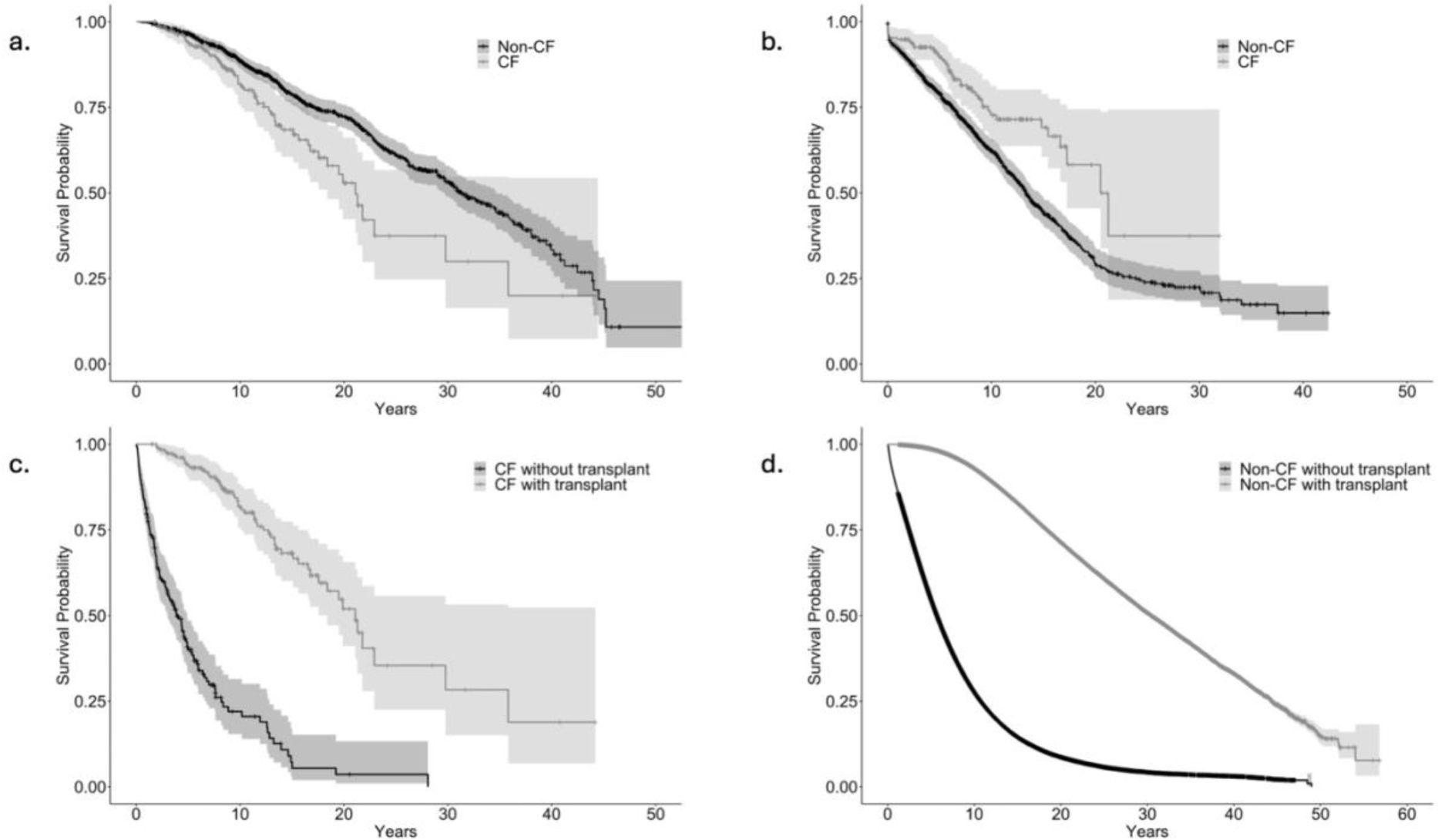
Patient and graft survival. a. Overall survival of ESKD transplant patients with CF, median 21.1 years, and without CF, median 31.8 years, (p = 0.00056), with propensity score matching for age at first ESKD service, BMI, Black and White race, Hispanic ethnicity, diabetes diagnosis, diabetes as cause of ESKD, lung transplant status, and complications of transplanted lung as primary cause of ESKD. b. Death-censored survival of first graft for patients with CF, median 20.5 years, and without CF, median 13.3 years (p < 0.0001), with propensity score matching for age at first ESKD service, BMI, Black and White race, Hispanic ethnicity, diabetes diagnosis, diabetes as cause of ESKD, lung transplant status, and complications of transplanted lung as primary cause of ESKD. c. Overall survival of CF patients with a kidney transplant, median 21.1 years, and without a kidney transplant, median 3.89 years (p < 0.0001) with propensity score matching for age at first ESKD service, BMI, Black and White race, Hispanic ethnicity, diabetes diagnosis, diabetes as cause of ESKD, lung transplant status, and complications of transplanted lung as primary cause of ESKD. d. Overall survival of patients without CF with a kidney transplant, median 30.5 years, and patients without a kidney transplant, median 5.69 years (p < 0.0001), with propensity score matching for age at first ESKD service, BMI, Black and White race, Hispanic ethnicity, diabetes diagnosis, diabetes as cause of ESKD, lung transplant status, and complications of transplanted lung as primary cause of ESKD.

#### Characteristics of individuals with CF with and without kidney transplant Demographic and characteristics

Comparing persons with CF who did and did not have a kidney transplant, those with a kidney transplant were significantly younger at ESKD incidence than those who did not receive a kidney transplant, 36.4 vs 47.6 years (p < 0.001), and had non-significantly higher odds of being female, 52.1% vs 50.3% (OR 1.25, 0.81, 1.93; p = 0.31) (**Table 2**). Those with a kidney transplant were non-significantly more likely to be identified as White, 91.7% vs 82.9% (OR 1.86, 0.94, 3.66; p = 0.07), or of Hispanic ethnicity 8.9% vs 7.0% (OR 1.13, 0.51, 2.52; p = 0.77), and had lower odds of being identified as Black or African American than those without, 6.3% vs 15.5% (OR 0.43; 0.20, 0.90; p = 0.03). Those with a kidney transplant had higher odds of nephrologist care prior to ESKD than those without (OR 1.90; 1.03, 3.53; p = 0.04).

**Table 2:** Characteristics of US patients with end stage kidney disease and cystic fibrosis with and without a kidney transplant.

|  | Transplant<br>(N=192) | Non-Transplant<br>(N=187) | Univariable |  |  | Age-adjusted<sup>1</sup> |  |  |
| --- | --- | --- | --- | --- | --- | --- | --- | --- |
|  |  |  | Coeff/OR<sup>2</sup> | 95% CI | p-value | Coeff/OR | 95% CI | p-value |
| **Female sex** | 100<br>(52.1%) | 94<br>(50.3%) | 1.08 | (0.72, 1.61) | 0.72 | 1.25 | (0.81, 1.93) | 0.31 |
| **Age at first ESKD service (Mean ± SD)** | 36.4 ± 14.2 | 47.6<br>(16.6) | -11.3 | (-14.4, -8.14) | <0.001 | - | - | - |
| **BMI (Mean ± SD)** | 21.8 ± 6.38 | 24.3 ± 7.19 | -2.41 | (-3.82, -1.00) | <0.001 | -1.16 | (-2.63, 0.30) | 0.12 |
| **Race** |  |  |  |  |  |  |  |  |
| Black/African American | 12<br>(6.3%) | 29<br>(15.5%) | 0.36 | (0.18, 0.74) | 0.005 | 0.43 | (0.20, 0.90) | 0.03 |
| White | 176<br>(91.7%) | 155<br>(82.9%) | 2.27 | (1.20, 4.30) | 0.01 | 1.86 | (0.94, 3.66) | 0.07 |
| **Hispanic ethnicity** | 17<br>(8.9%) | 13<br>(7.0%) | 1.30 | (0.61, 2.76) | 0.49 | 1.13 | (0.51, 2.51) | 0.77 |
| **ESKD Primary Cause<sup>3</sup>** |  |  |  |  |  |  |  |  |
| Diabetes | 28<br>(14.6%) | 64<br>(34.2%) | 0.33 | (0.20, 0.54) | <0.001 | 0.39 | (0.23, 0.67) | <0.001 |
| Type 1 | 13<br>(46.4%) | 19<br>(29.7%) | 0.64 | (0.31, 1.34) | 0.24 | 0.62 | (0.28, 1.35) | 0.23 |
| Type 2 | 15<br>(53.6%) | 45<br>(70.3%) | 0.27 | (0.14, 0.50) | <0.001 | 0.35 | (0.18, 0.68) | 0.002 |
| Complications of transplanted lung | 30<br>(15.6%) | 34<br>(18.2%) | 0.83 | (0.49, 1.43) | 0.51 | 0.75 | (0.42, 1.32) | 0.32 |
| Complications of any transplanted organ | 42<br>(21.9%) | 36<br>(19.3%) | 1.17 | (0.71, 1.93) | 0.53 | 1.06 | (0.62, 1.80) | 0.83 |
| Nephropathy due to toxins and medications<sup>4</sup> | 24<br>(12.5%) | ≤10 | - | - | - | - | - | - |
| Hypertension<sup>4</sup> | ≤10 | 28<br>(15.0%) | - | - | - | - | - | - |
| Tubular necrosis<sup>4</sup> | ≤10 | 11<br>(5.9%) | - | - | - | - | - | - |
| Glomerulonephritis | 23<br>(12.0%) | 14<br>(7.5%) | 1.68 | (0.84, 3.38) | 0.14 | 1.37 | (0.66, 2.86) | 0.40 |
| **Comorbidities** |  |  |  |  |  |  |  |  |
| Diabetes diagnosis | 118<br>(61.5%) | 123<br>(65.8%) | 0.83 | (0.55, 1.26) | 0.38 | 0.95 | (0.61, 1.49) | 0.83 |
| Lung transplant status | 123<br>(64.1%) | 68<br>(36.4%) | 3.12 | (2.05, 4.74) | <0.001 | 2.63 | (1.70, 4.09) | 0.02 |
| Liver transplant status<sup>4</sup> | 22<br>(11.5%) | ≤10 | - | - | - | - | - | - |
| Pancreas transplant status<sup>4</sup> | 22<br>(11.5%) | ≤10 | - | - | - | - | - | - |
| **Pre-ESKD care by a nephrologist** | 111<br>(57.8%) | 115<br>(61.5%) | 1.84 | (1.02, 3.31) | 0.04 | 1.90 | (1.03, 3.53) | 0.04 |
| **Death<sup>5</sup>** | 45<br>(23.4%) | 117<br>(62.6%) | 0.18 | (0.12, 0.29) | <0.001 | 0.21 | (0.13, 0.33) | <0.001 |

<sup>1</sup>Adjusted for age at first ESKD service, <sup>2</sup>Linear regression coefficients are given for age at first ESKD service, BMI, and years in dialysis. Odds ratios are derived by logistic regression. For all ORs, non-CF patients were used as the reference group. <sup>3</sup>Not all possible primary cause categories were included in this analysis, <sup>4</sup>All calculations involving cells with fewer than 11 individuals were censored (-), <sup>5</sup>Death prior to December 31, 2018.

#### Primary causes of ESKD

Those with a kidney transplant had lower odds of diabetes mellitus being the primary cause of ESKD, 14.6% vs 34.2% (OR 0.39; 0.23, 0.67; p < 0.001). They had non-significantly lower odds than those without a kidney transplant of complications of lung transplantation being the primary cause of ESKD, 15.6% vs 18.2% (OR 0.75; 0.42, 1.32; p = 0.32) and non-significantly higher odds of complications of lung or other organ being the primary cause of ESKD, 21.9% vs 19.3% (OR 1.06; 0.62, 1.80; p = 0.83). Those with kidney transplant had higher odds of nephropathy due to toxins or medications being the primary cause of ESKD, 12.5% vs 3.7% (OR 3.36; 1.36, 8.33; p = 0.009).

#### Mortality and survival

In age-adjusted univariable analysis, those with CF with a kidney transplant had lower odds of death prior to 31 December 2018 than those who did not have a kidney transplant, 23.4% vs 62.6% (OR 0.21; 0.13, 0.33; p < 0.001). In Kaplan-Meier analysis with propensity score matching, median patient survival was 21.1 years for those with CF with a kidney transplant, and 3.9 years for those with CF without a kidney transplant (p < 0.0001) (Figure 1c). In Kaplan-Meier analysis with propensity score matching comparing all without CF, median patient survival was 30.5 years with a kidney transplant and 5.7 years without a kidney transplant (Figure 1d).

## Discussion

CF is widely understood to be a devastating chronic condition with features which may discourage referral and listing for kidney transplantation. Life expectancy and general health in people with CF has improved markedly in recent decades due to advances in prevention and management of infection and other complications, with predicted median survival for a person with CF born today in the US estimated at 53.1 years^8^. Further improvements in health, life expectancy, and quality of life are anticipated following FDA approval of highly effective modulators of the CFTR in 2019^9,10^. Previous investigators have documented an increased risk of CKD in people with CF^4,11,12^, which may progress to ESKD, raising the question of suitability or otherwise of these individuals for referral and evaluation for kidney transplantation. The demographics, disease characteristics, and patient and graft outcomes following kidney transplant in persons with CF have not previously been described. People with CF who underwent kidney transplantation were younger than both those with CF who did not receive a transplant and those with a transplant who did not have CF but were considerably older than the general population with CF in 2016, whose mean age was 19 years. This suggests that persons with CF who develop ESKD may have traits which increase susceptibility to kidney disease and specific characteristics promoting longevity with CF. Those with CF who underwent kidney transplantation were more likely to be female, in contrast to the equal sex distribution of CF in the US^8^. As previously documented in the general ESKD population, persons with CF who were identified as Black or African American had lower likelihood of receiving a kidney transplant relative to their representation in the CF population with ESKD. This disparity is likely to be due to multiple factors influencing referral and listing for kidney transplant in diverse populations^13^.

The spectrum of causes of ESKD in those with CF was quite different from that of the general ESKD population. Diabetes mellitus was less frequently the etiology of ESKD in those with CF than in those without CF and was also less frequent in those with CF who received a transplant than in those with CF who did not. Cystic fibrosis-related diabetes (CFRD) is a specific non-autoimmune entity with features of both type 1 and type 2 diabetes and is a consequence of pancreatic inflammation and fibrosis. The rate of CFRD increases with age, with lifetime incidence of 50-80%^14^. Microvascular complications, including diabetic nephropathy, retinopathy, and hypertension occur in CFRD^15^. However, the rate of diabetic nephropathy leading to ESKD, and the risk of recurrence of diabetic nephropathy following kidney transplant in those with CFRD are not known and are areas of interest for future investigation. Type 1 and type 2 endemic diabetes mellitus can also occur in people with CF. We did not distinguish CFRD from type 1 or type 2 diabetes in this study because CFRD is not a specified diagnosis on the ESKD Medical Evidence Report. However, we infer that at least a proportion had CFRD, because CFRD is insulin-dependent in later stages, so more likely to be classified as type 1, and a higher proportion of those with CF than those without CF were classified as type 1.

Organ transplantation, particularly of lung, was the second most frequent primary cause of ESKD in those with CF who underwent kidney transplantation. A striking finding was that lung transplant status was documented in two thirds of those with CF, and in only 0.2% of those who did not have CF. Pancreas and liver transplant were also more frequent, both as diagnoses and as primary etiology of ESKD. Lung transplant is employed as a last resort therapy in individuals who have suffered repeated pulmonary exacerbations, generally requiring treatment with nephrotoxic antibiotics including aminoglycosides, vancomycin, and amikacin^16^. Repeated aminoglycoside exposure is likely to contribute to the risk of CKD associated with lung transplant, although the role of aminoglycosides in CKD in CF has been controversial^17–19^. Renal complications are frequent following lung transplantation surgery^20^, and kidney function declines more rapidly following lung transplantation for CF than those transplanted for other reasons^21^. Acute kidney injury (AKI), which predisposes to CKD^22^, occurs in the perioperative period in half of all lung transplant patients, nearly one in ten of whom require dialysis^20^. In addition, subsequent immunosuppression with calcineurin inhibitors, together with increased tendency to dehydration due to obligatory solute loss in lungs and the gastrointestinal (GI) tract, and CF-specific renal tubular functional abnormalities^1,23,24^, are all likely to contribute to the increased risk of cumulative nephron loss following lung transplantation in CF. Finally, patient selection for kidney transplant may be biased towards those with CF and lung, liver, or pancreas transplant because these patients are likely to be followed in multidisciplinary transplant centers with experience in kidney transplantation in CF.

People with CF had shorter time on dialysis prior to transplant, and were significantly more likely to receive a living donor graft than those with ESKD who did not have CF. This may speak to the close involvement of patient, family, and provider support networks involved in the CF community. Those with CF who received a kidney transplant were more likely than those who did not to have received nephrology care prior to ESKD incidence, suggesting that access to appropriate health care services is relevant to the odds of transplantation for persons with CF. Perhaps the most counterintuitive and consequential findings of this study were that graft and patient survival were of similar magnitude to that experienced by people without CF. In fact, death-censored graft survival in those with CF was significantly longer than that of those without CF. Overall median patient survival following the first kidney transplant with CF was 21.1 years. While this is shorter than the survival period for matched patients without CF, it is substantial, and represents a comparable survival benefit when compared to dialysis. It is our hope that these findings will both increase the awareness of kidney disease in the CF community and encourage early referral and positive consideration for kidney transplant in this unique population.

These data provide a baseline for assessing the future impact of newer therapies on those with CF and ESKD who may be eligible for kidney transplantation. Highly effective modulators of the CFTR were approved for use in the US in 2019, and progression of kidney disease to ESKD typically takes several years. These agents would have been highly unlikely to have impacted the characteristics or outcomes of kidney transplantation during the period of data collection for this study. Highly effective modulators of the CFTR may both prolong life and preserve renal function^7,8^. Their impact on kidney disease in CF, and that of possible future gene-editing therapies is an area of great future interest.

## Limitations of this study

This is a retrospective observational study using de-identified data. Cystic fibrosis is not a specified diagnosis on the ESKD intake form. As detailed in the methods, we relied on comparisons between individuals in the USRDS Hospitalized data set with and without a diagnosis of CF.

## Disclaimer

The data reported here have been supplied by the United States Renal Data System (USRDS). The interpretation and reporting of these data are the responsibility of the author(s) and in no way should be seen as an official policy or interpretation of the U.S. government.

## Data Availability

All data produced in the present study are available upon reasonable request to the authors

